# Multilevel determinants of Covid-19 vaccine hesitancy and undervaccination among marginalized populations in the United States: A scoping review

**DOI:** 10.1101/2023.02.23.23286342

**Authors:** Peter A. Newman, Thabani Nyoni, Kate Allan, Sophia Fantus, Duy Dinh, Suchon Tepjan, Luke Reid, Adrian Guta

## Abstract

**Background:** Amid persistent disparities in Covid-19 vaccination, we conducted a scoping review to identify multilevel determinants of Covid-19 vaccine hesitancy (VH) and undervaccination among marginalized populations in the U.S.

**Methods:** We utilized the scoping review methodology developed by the Joanna Briggs Institute and report all findings according to PRISMA-ScR guidelines. We developed a search string and explored 7 databases to identify peer-reviewed articles published from January 1, 2020–October 31, 2021, the initial period of U.S. Covid-19 vaccine avails.comability. We combine frequency analysis and narrative synthesis to describe factors influencing Covid-19 vaccination among marginalized populations.

**Results:** The search captured 2,496 non-duplicated records, which were scoped to 50 peer-reviewed articles: 11 (22%) focused on African American/Black people, 9 (18%) people with disabilities, 4 (8%) justice-involved people, and 2 (4%) each on Latinx, people living with HIV/AIDS, people who use drugs, and LGBTQ+ people. Forty-four articles identified structural factors, 36 social/community, 27 individual, and 40 vaccine-specific factors. Structural factors comprised medical mistrust (of healthcare systems, government public health) and access barriers due to unemployment, unstable housing, lack of transportation, no/low paid sick days, low internet/digital technology access, and lack of culturally and linguistically appropriate information. Social/community factors including trust in a personal healthcare provider (HCP), altruism, family influence, and social proofing mitigated VH. At the individual level, low perceived Covid-19 threat and negative vaccine attitudes were associated with VH.

**Discussion:** This review indicates the importance of identifying and disaggregating structural factors underlying Covid-19 undervaccination among marginalized populations, both cross-cutting and population-specific—including multiple logistical and economic barriers in access, and systemic mistrust of healthcare systems and government public health—from individual and social/community factors, including trust in personal HCPs/clinics as reliable sources of vaccine information, altruistic motivations, and family influence, to effectively address individual decisional conflict underlying VH as well as broader determinants of undervaccination.

## Introduction

Vaccination remains an essential component of public health strategies to control the Covid-19 pandemic. Vaccination has reduced Covid-19-related morbidity and mortality in the United States (U.S.) [1], and globally—with an estimated 63% (14.4–19.8 million) reduction in deaths in the first year of Covid-19 vaccination alone [2]. Vaccine booster doses [3], and bivalent vaccine boosters compared with past monovalent vaccination [4], have further reduced Covid-19-associated hospitalization and death. Yet, despite the tremendous successes of Covid-19 vaccination, pervasive challenges ascribed to vaccine hesitancy (VH), and broader Covid-19 undervaccination, threaten the effectiveness of vaccines in controlling Covid-19 and other vaccine-preventable diseases.

The World Health Organization (WHO) Strategic Advisory Group of Experts (SAGE) on Immunization defines vaccine hesitancy (VH) as “delay in acceptance or refusal of vaccination despite availability of vaccination services” [5,p.4161]. In addition to individual-level correlates of VH, SAGE [6] along with other research initiatives [7,8] have drawn attention to the critical importance of vaccine availability and access—amid systematic variations in vaccine coverage across countries and subpopulations—and issues that may pertain to specific vaccines. Collectively, these challenges support the need to identify and examine multilevel factors that contribute to Covid-19 VH and broader Covid-19 undervaccination (e.g., due to inaccessibility, unavailability) [9].

Disparities in Covid-19 morbidity and mortality, as well as lower rates of Covid-19 vaccination, are well documented among marginalized populations in the U.S. [10-13]. Marginalization is defined by systemic processes driven by ideologies such as racism, sexism, heterosexism, cissexism, ableism, and xenophobia, that exclude certain populations or relegate them to the periphery of political and socioeconomic resources [14,15]. The structural and social inequities produced by marginalization create adverse social determinants of health (SDOH)— such as residential segregation and housing insecurity, employment precarity, income insecurity, lack of access to affordable healthcare, barriers to physical mobility, and stigmatization—all of which increase vulnerability to poor health outcomes [14]. Adverse SDOH exacerbate vulnerability to Covid-19 through tthe disproportionate prevalence of underlying conditions associated with worse Covid-19 outcomes, lower levels of health insurance and healthcare access, and by impeding the ability to practice public health measures to reduce viral transmission [16-19]. Although heightened vulnerability indicates an increased need for vaccination, these same SDOH may contribute to *lower* levels of Covid-19 vaccination [20,21]. In fact, across 45 U.S. states, 0.1%–24.9% lower vaccination was documented among Blacks vs. Whites, 0.0%–37.6% lower among Hispanics vs. Whites, as of May 2021—with state-level rate differences significantly associated with a metric of structural racism [21]. In a national study, eligible healthcare facilities and community pharmacies in counties with higher Black composition were significantly less likely to provide Covid-19 vaccination [20].

Although some of the racial/ethnic disparities in Covid-19 vaccination documented earlier in the pandemic have reportedly narrowed over time, attributed in part to the success of targeted interventions [11], disparities persist in lower administration of booster doses among Black, Hispanic/Latinx, and American Indian populations versus Whites [22]. Consistent with the “inverse equity hypothesis,” with each innovation in Covid-19 vaccines, uptake is predicted to be greater among wealthier and more socially connected segments of the population, thereby increasing rather than reducing inequity [23,24]. Nevertheless, population-level disparities in Covid-19 vaccination are often conceptualized as contingent on individual-level factors (e.g., low perceived threat, vaccine attitudes) to the relative underemphasis of structural and systemic factors [20,21,25,26]. Moreover, trust/mistrust in a personal HCP as a source of reliable information [27] is often conflated with systemic medical mistrust: distrust of healthcare systems, government public health, medical treatments, and/or the pharmaceutical industry arising from present-day experiences of discrimination and/or historical experiences of discrimination and injustice in the healthcare system [28-30].

Building on the SAGE “Vaccine Hesitancy Determinants Matrix” and recommendations to approach VH as a population- and context-specific phenomenon [6,31], along with critiques of the lack of clarity in the definition and application of VH [32-34], we conducted a scoping review to examine multilevel factors associated with Covid-19 VH and broader Covid-19 undervaccination among marginalized populations in the U.S.

## Methods

A scoping review was conducted given our aim of mapping the evidence landscape [35] on factors associated with Covid-19 VH and undervaccination among marginalized populations. A scoping review protocol, including our search strategy and review process, has been previously published [36]. We utilized the scoping review methodology described by Arksey and O’Malley [37], further developed by the Joanna Briggs Institute [38], with all results reported in accordance with PRISMA Extension for Scoping Reviews (PRISMA-ScR) guidelines [39].

### Research question

This scoping review was guided by the question, “What are the determinants of Covid-19 ‘vaccine hesitancy’ reported among marginalized populations in the U.S. and Canada?”, with sub-questions, “what are the focal populations in studies of Covid-19 VH and undervaccination among marginalized populations?” and “what are the contexts and determinants of Covid-19 VH and undervaccination by structural, social and community, individual, and vaccine-specific factors?” [36].

### Information sources and search strategy

We identified the following list of databases to search in consultation with a specialist research librarian: Medline, Embase, Cochrane CENTRAL, Cochrane Covid-19 Study Register, PsychINFO, Sociological Abstracts, and the International Bibliography of the Social Sciences (IBSS). We then developed a search string with keywords and synonyms related to vaccine hesitancy (i.e., “vaccine hesitancy”, “vaccine refusal”, “vaccine confidence”, “vaccine distrust”, “vaccine mistrust”, “vaccine barriers”) and Covid-19 (i.e., “corona virus” or “coronavirus” or “COVID” or “nCoV” along with adjacent terms “19” or “2019, or “SARS-CoV-2” or “SARS Coronavirus 2”, etc.). S1 Table shows an example search string for Medline and Embase. We adapted the search string to accommodate differences in syntax and parameters among the selected databases.

### Study selection criteria

We developed inclusion and exclusion criteria prior to conducting the search for peer-reviewed articles published between January 1, 2020 and October 31, 2021: 1) populations in the U.S. or Canada; 2) adults ≥18 years; 3) focused on (>50%) marginalized populations, 4) focused on Covid-19 vaccine hesitancy, refusal, confidence, mistrust, or barriers. Articles were excluded if they were 1) published prior to 2020; 2) not written in English; 3) focused on healthcare workers, staff, or students/interns; or 4) focused on children (<18 years) or parents’ vaccination of children. We selected search dates corresponding to the declaration of Covid-19 as a public health emergency through the initial authorization of Covid-19 vaccines for the adult population in the U.S. and Canada in December 2020, until U.S. FDA and Health Canada recommendations for the first booster vaccines, beginning in late September 2021 to early November 2021 [40,41].

We identified marginalized populations informed by a review of populations particularly vulnerable to discrimination and exclusion in healthcare [42] or due to broader adverse SDOH [14]. The populations include those marginalized in relation to their race or ethnicity, sexual orientation or gender identity, physical or mental disability, persons living with HIV, immigrants/refugees, persons who experience homelessness, justice-involved people, and people who use drugs, along with their intersections (e.g., Latinx immigrants).

### Study selection process

Search results of peer-reviewed articles were uploaded into Covidence software. Multiple research team discussions ensured consistent application of inclusion/exclusion criteria. Initially groups of two reviewers (among LR, TN, DD, KA, SF or ST) independently screened abstracts and titles until we attained inter-rater reliability of 90% [43]; this was achieved after two rounds of abstract review and team discussions to achieve consensus. Subsequently, in accordance with rapid review methods, one reviewer screened each abstract/title to determine inclusion or exclusion [44]. One reviewer (TN, DD, KA, SF or ST) then screened each full-text for inclusion. Full-text articles designated for exclusion were then reviewed by a single arbitrator (PN), with discrepancies resolved by consensus among the research team [44,45].

### Data extraction and synthesis

We abstracted data on publication characteristics (i.e., author(s), study sites, sample size, populations, methods, and domains of VH factors identified (see Table 1). We then reviewed the abstracted data using quantitative (i.e., frequency) analysis to describe study locales, sample sizes, focal populations, methods, and VH factor domains, and qualitative (i.e., narrative) analysis to synthesize the evidence on VH and undervaccination among each subpopulation.

**Table 1.**
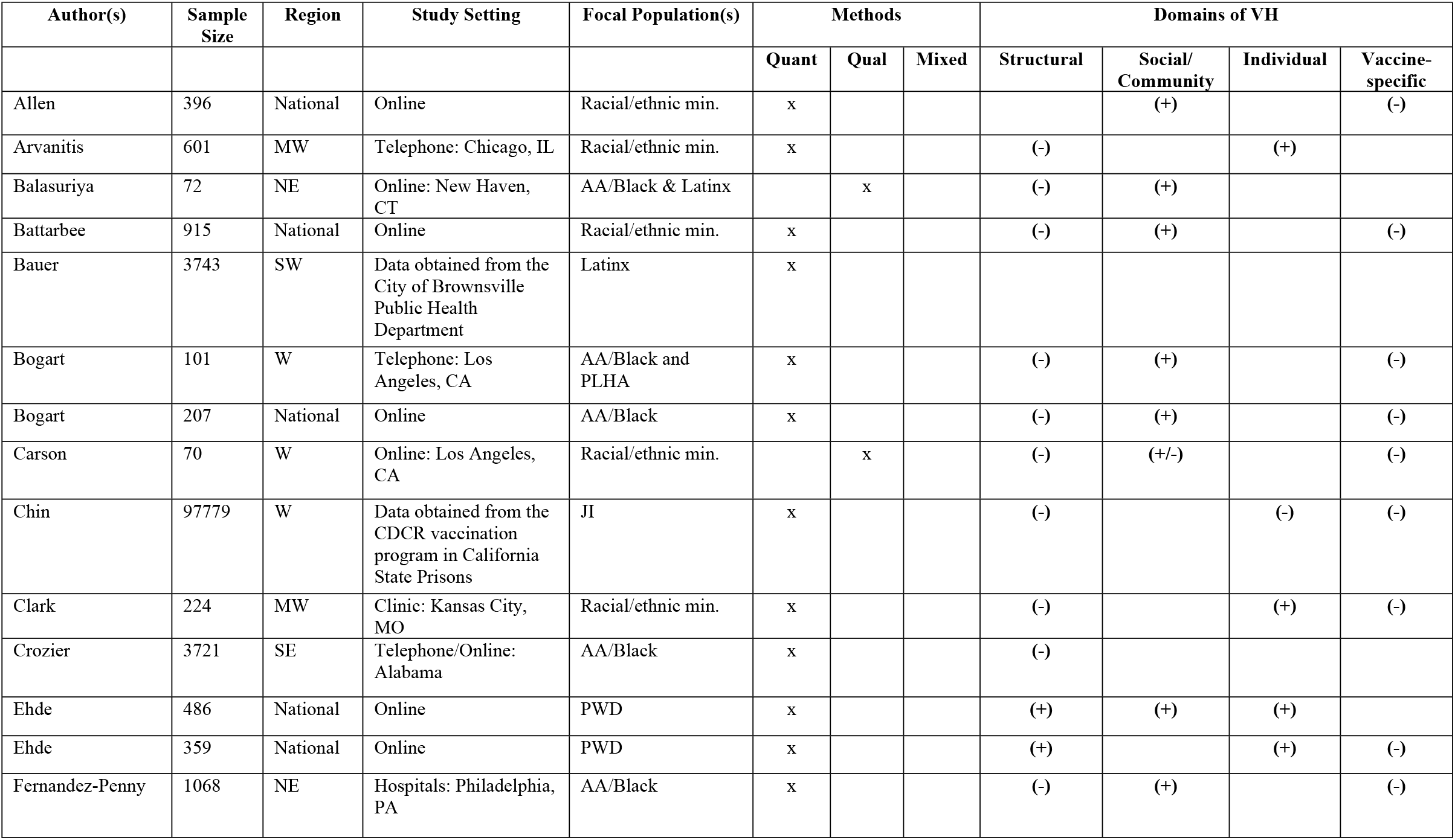
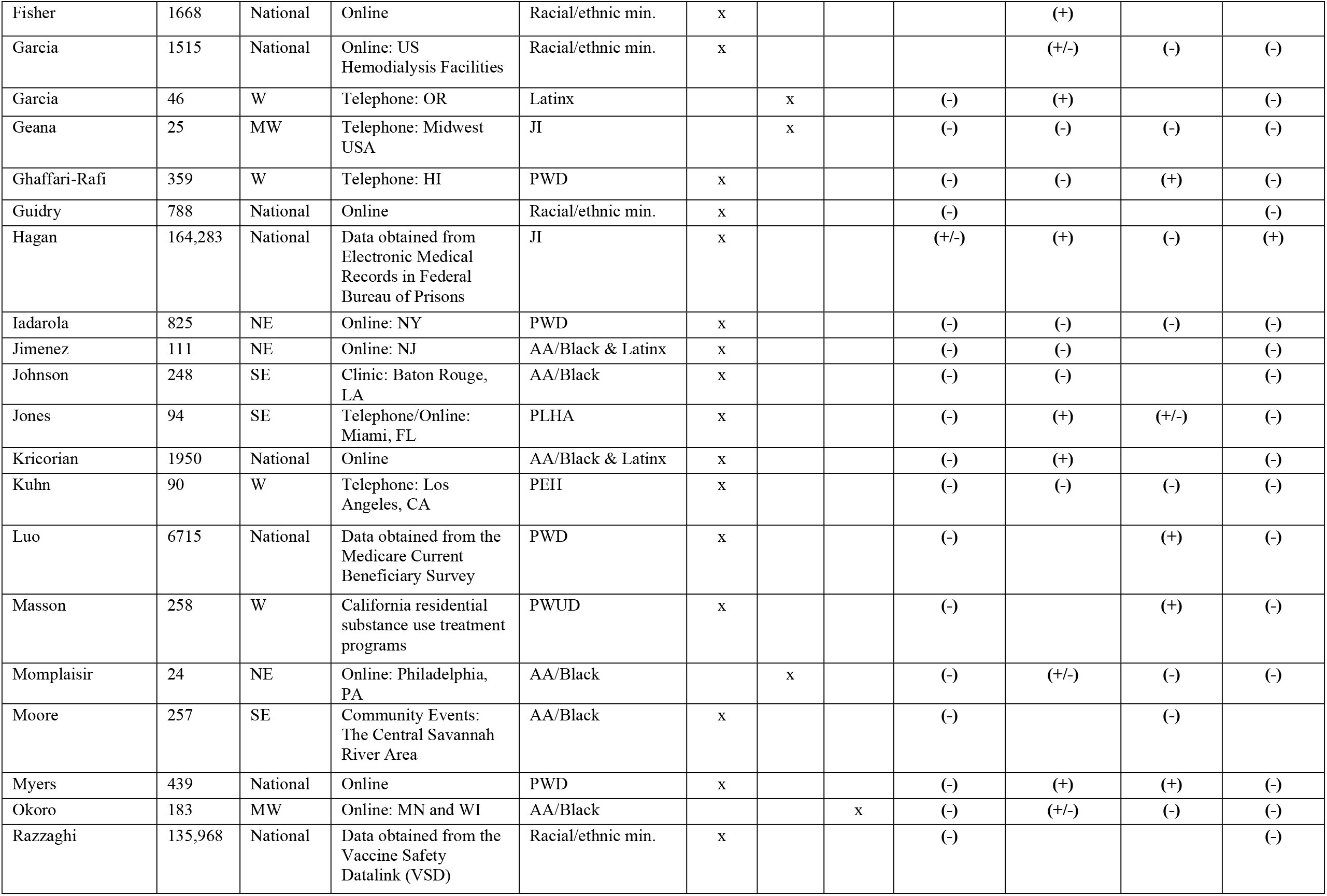
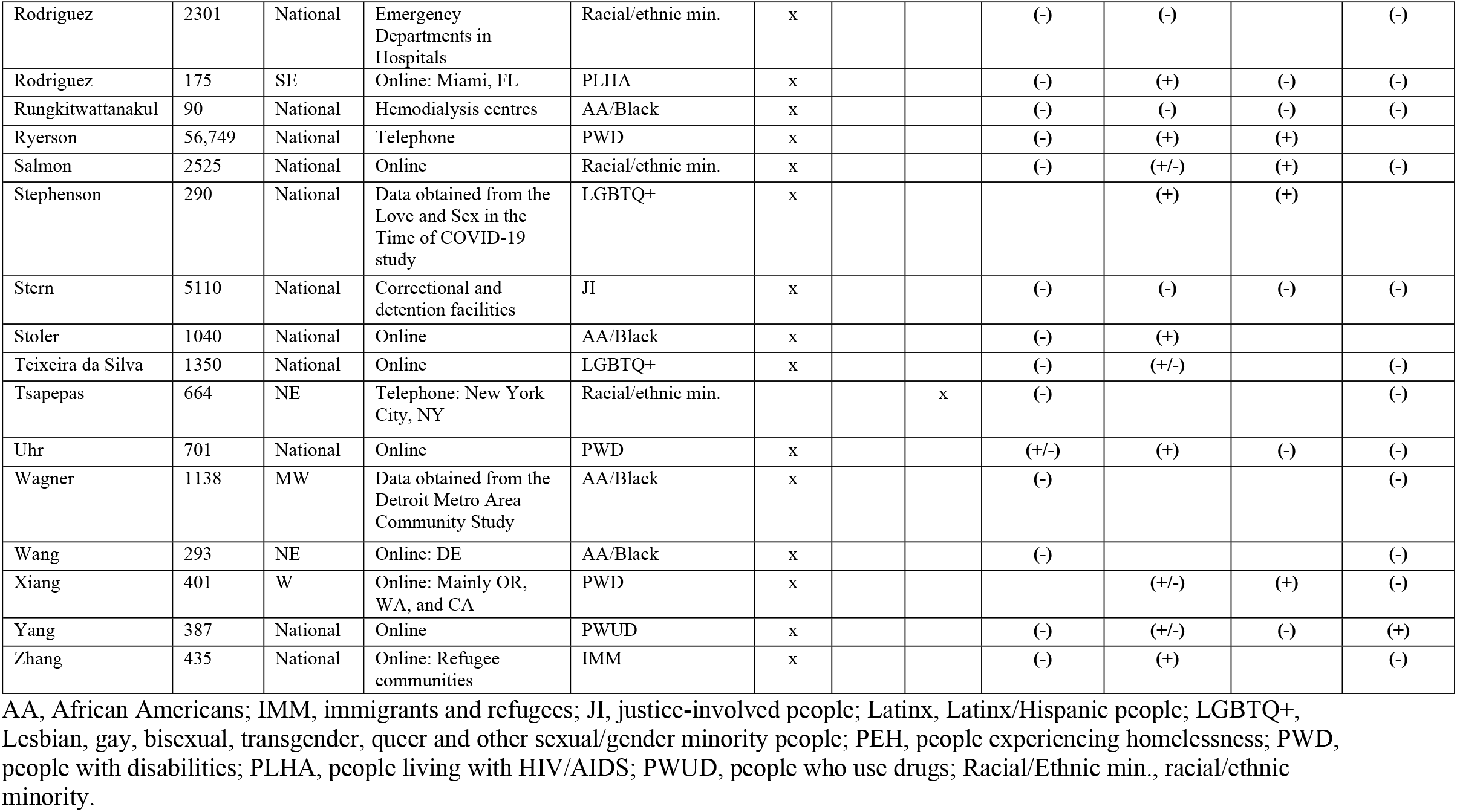
Study characteristics, focal populations, and domains of Covid-19 vaccine hesitancy and undervaccination among marginalized populations in the U.S., January 1, 2020 to October 31, 2021 (n = 50 articles)

All authors categorized publications by domains of factors identified, using a four-component framework informed by the “Vaccine Hesitancy Determinants Matrix” [6,31] and recommendations to disaggregate multilevel phenomena [8,33,34]. Domains comprised structural (e.g., access to vaccination site, medical mistrust); social/community (e.g., family support, trust in personal HCP or clinic, social proofing [i.e., based on actions of others in one’s community], community benefits); individual (e.g., perceived Covid-19 threat, perceived transmission risk, general vaccine beliefs); and Covid-19 vaccine-specific factors (e.g., vaccine safety, side effects, efficacy). One change to data extraction (but not inclusion) was made from the initial protocol in that we differentiated between medical mistrust at the “system-level” and trust in a personal HCP/clinic.

## Results

The search captured 4,399 peer-reviewed articles in total, with 2,496 abstracts screened after removal of duplicates. We identified 363 full-text articles to be screened for eligibility, with a total of 50 peer-reviewed articles included [12,26,46-93]. The PRISMA flow diagram shows the process of identifying relevant peer-reviewed journal articles (see Fig 1).

**Figure 1.**
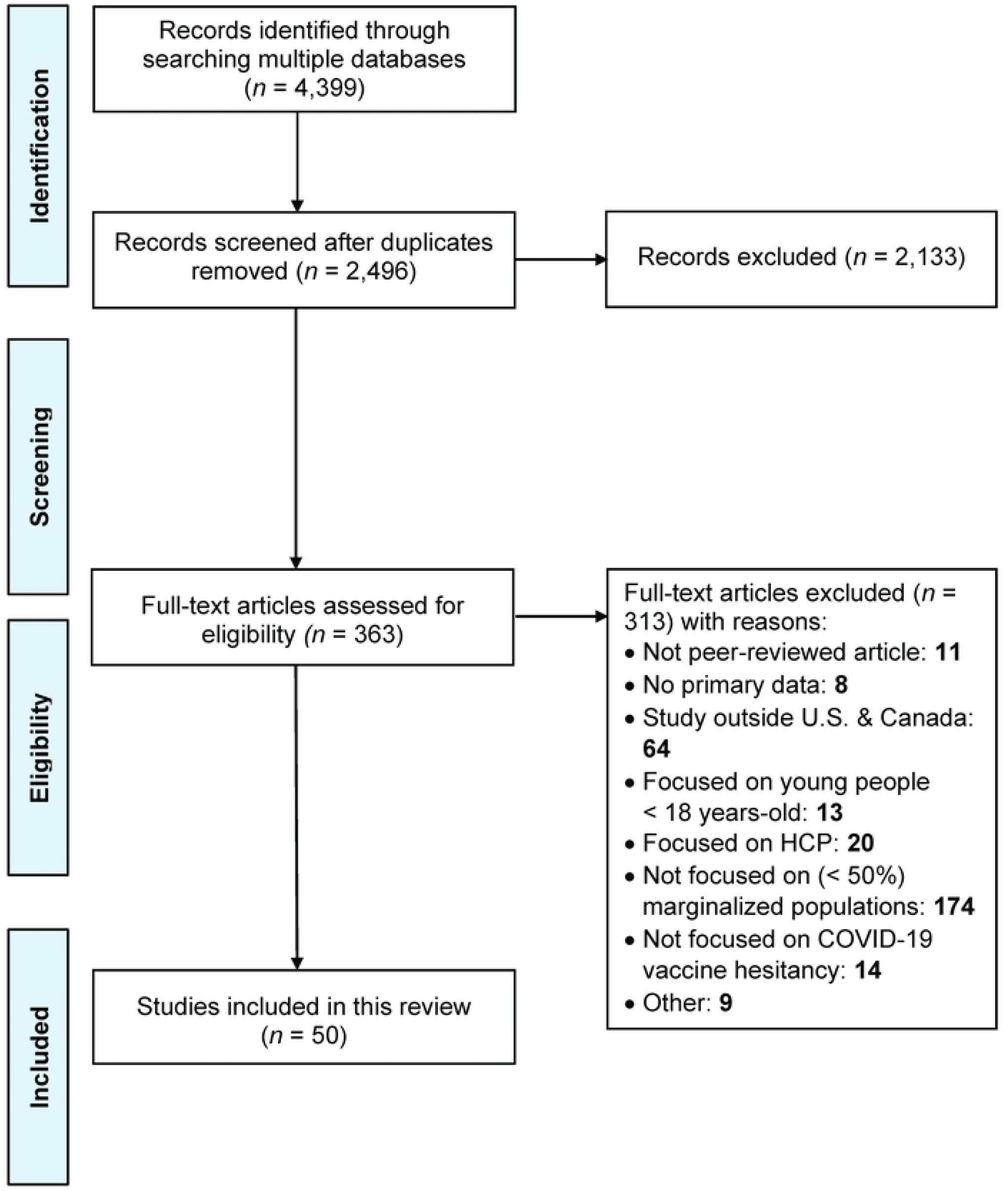
PRISMA flow diagram for scoping review of COVID-19 vaccine hesitancy and undervaccination among marginalized populations.

### Description of studies

Among the 50 articles included, 86% were quantitative studies, 10% qualitative, and 4% mixed methods (see Table 1). Sample sizes ranged from n=24 to n=164,283 (N=499,237; median=437). All studies were conducted in the U.S., including 24 (48%) national studies, 8 (16%) in the West, 7 (14%) Northeast, 5 (10%) Midwest, 5 (10%) Southeast, and 1 (2%) in the Southwest region. No studies were identified from Canada that met inclusion criteria within the timeframe of the review.

Table 2 shows the number of focal articles by subpopulation, with 11 (22%) focused on African American/Black people, 9 (18%) on people with disabilities, 4 (8%) on justice-involved people, and 2 (4%) each on Latinx, people living with HIV/AIDS (PLHA), people who use drugs (PWUD), and lesbian, gay, bisexual, transgender, and queer (LGBTQ+) people. We allocated findings from articles broadly focused on racial/ethnic minorities to each of the subpopulations (i.e., African American/Black, Latinx, Asian, American Indian) for which disaggregated data were provided (i.e., one article could be a “source” for several populations). Four additional articles addressing VH pertaining to two subpopulations (e.g., African American/Black PLHA) were each allocated as two sources. Table 2 shows the total number of sources (n=73) addressing population-specific factors associated with VH across the 50 articles. Among these, 41.1% of sources (n=30) identified VH factors among African American/Black populations, 17.8% (n=13) Latinx populations, 12.3% (n=9) people with disabilities, and 5.5% (n=4) each Asian, justice-involved, and PLHA.

**Table 2.** Focal articles and additional sources by subpopulation.

| **Subpopulation** | **Focal articles on each sub-population** | **Additional sources addressing each subpopulation** | **Number (%) of total sources by sub-population (n=73)** |
| --- | --- | --- | --- |
| African American/Black | 11 | 19 | 30 (41) |
| Latinx | 2 | 11 | 13 (18) |
| People with disabilities | 9 | 0 | 9 (12) |
| Justice involved | 4 | 0 | 4 (5) |
| People living with HIV/AIDS | 2 | 2 | 4 (5) |
| Asian people | 0 | 4 | 4 (5) |
| People who use drugs | 2 | 1 | 3 (4) |
| Immigrants/refugees | 1 | 1 | 2 (3) |
| LGBTQ+ people | 2 | 0 | 2 (3) |
| People experiencing homelessness | 1 | 0 | 1 (1) |
| American Indians/Native | 0 | 1 | 1 (1) |
| **Subtotal** | **34** | **39** | **73** |

LGBTQ+, Lesbian, gay, bisexual, transgender, queer and other sexual/gender minority people

Fig 2 shows the number of articles addressing factors associated with VH in each of four domains: structural, social/community, individual, and Covid-19-vaccine-specific. Table 3 shows the domains of factors associated with VH in each subpopulation. Next, we describe findings based on a narrative synthesis of factors associated with VH and undervaccination for each focal population.

**Fig 2.**
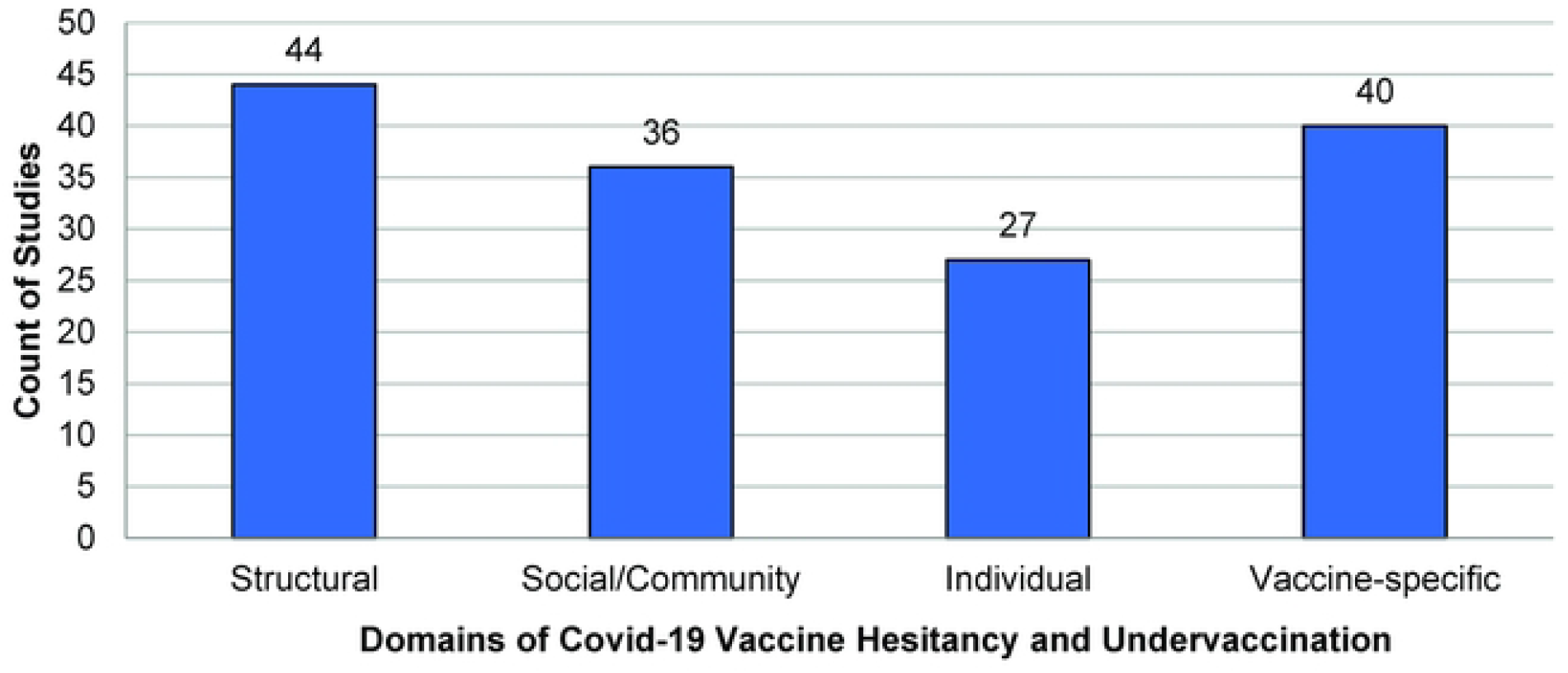
Count of articles identifying factors in each of the four domains of Covid-19 vaccine hesitancy and underutilization.

**Table 3.**
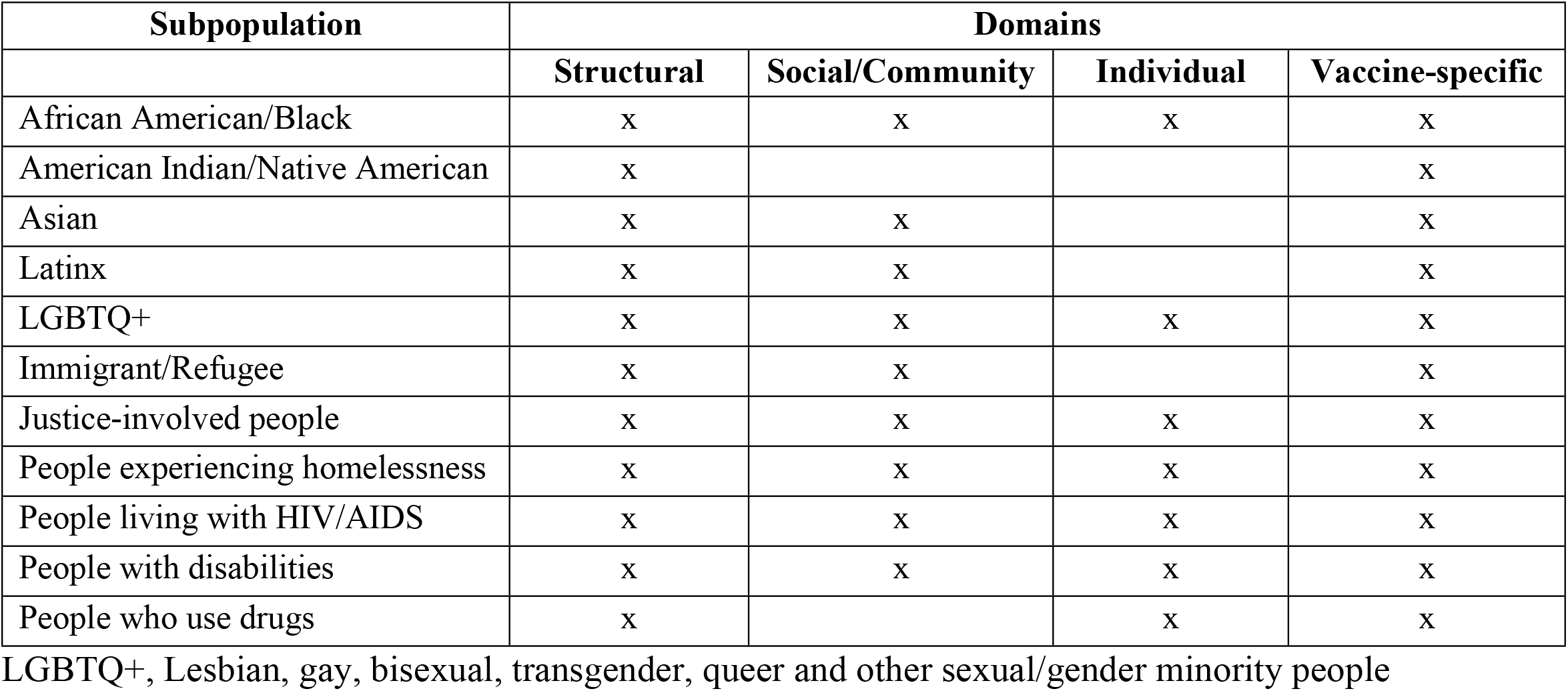
Population by domains of factors identified in Covid-19 vaccine hesitancy and undervaccination.

#### Black and African American people

Eleven articles focused exclusively on Covid-19 vaccination among Black/African Americans [26,56,59,69,74,75,77,81,85,89,90], and 19 additional articles provided disaggregated data—a total of 30 sources [46-48,49,51-53,55,60,65-68,70,73,82,84,86,92]. At the structural level, 27 sources addressed the negative impacts of medical mistrust and distrust of government on Covid-19 vaccination among Black and African Americans [26,46-49,51-53,55,59,65-70,73-75,77,81,82,84-86,89,92]. Several studies described Covid-19 VH, such as due to fears of being experimented on with Covid-19 vaccines without consent, “based on…knowledge of historic mistreatment and unethical research practices that adversely impacted black patients and participants” [74, p.1786], including the legacy of the Tuskegee Study of untreated syphilis [53,67,68,77]. Eleven sources addressed barriers to present-day healthcare and Covid-19 vaccines for Black/African Americans, including unemployment, housing insecurity, limited interactions with HCP, and difficulties travelling to vaccination sites [48,52,53,56,68-70,75,77,89,90].

At the social/community level, nine sources indicated the role of trust in HCPs in Covid-19 vaccination intentions [26,46,51,60,69,70,74,82,85]. Characteristic of many studies, “Blacks and Hispanics were more likely than Whites to state that it was ‘extremely’ important that a medical professional of their same race/ethnicity endorse the vaccine before they took it” [26,p.8]. Seven studies addressed the influence of families and communities on Covid-19 vaccination [48,52,59,70,74,77,81], and further that “governmental agencies are less well-received as information sources compared to friends or family” [59, p.1103]. In one study, family influence promoted VH by casting doubt on public health information, rather than motivating vaccination [81].

At the individual level, four studies addressed negative impacts of religious beliefs, including preference for ‘natural’ medicines over vaccines, and general vaccine aversion on Covid-19 vaccination [67,74,77,81]. Covid-19 vaccine-specific concerns, identified in 14 studies, included vaccine safety, adverse effects, and uncertain efficacy [26,51-53,59,67-70,73,74,77,81,90], and the importance of evidence of Covid-19 vaccine safety and efficacy specific to Black/African Americans: “any data that’s connected with some of the conditions that are prevalent in our community” [53, p.8].

#### Latinx/Hispanic people

Two studies focused on Covid-19 vaccination among Latinx/Hispanic people [50,61], with an additional 11 sources providing data specific to this population [26,46,48,49,53,60,67,68,70,73,82]. At the structural level, five studies identified negative impacts of medical mistrust [48,49,61,67,82], including fear of being used as “guinea pigs (conejillos de indias)” and of being sterilized [61]. Limited access to information in one’s preferred language, complicated online vaccination sign-up procedures, lack of insurance, and lack of transportation to vaccination sites posed barriers to Covid-19 vaccination [48,53,61,68,70]. Two studies underscored the importance of tailored vaccine education and accessible information [68]: “[a family member] doesn’t speak English…so, a lot of the information that he may have heard about the vaccine and Covid probably came off of Univision and Telemundo” [48, p.6].

At the social/community level, five studies described the important role of personal HCPs in Covid-19 vaccination among Latinx communities [26,46,60,70,82], including endorsement from an HCP of one’s own ethnicity [26]. Two studies addressed altruism and wanting to protect one’s community as a motivator of Covid-19 vaccination [53,61]. Families and close communities were described as playing an important role in Covid-19 vaccination knowledge and decision-making [48,61,70]: “79% of the youth had discussed health with their families for an average of 50 minutes [per week]” [61, p.753]. No individual-level factors were described, but Covid-19 vaccine-specific concerns arose around safety, adverse effects, and efficacy [26,61,67,68,70,73].

#### Asian people

No studies focused exclusively on Covid-19 vaccination among people of Asian descent in the U.S.; four sources provided disaggregated data on Asian people [46,53,64,67]. At the structural level, barriers included difficulties in accessing reliable translations and messengers of Covid-19 vaccine information [53]. Beyond linguistic translation, a Filipinx participant explained, “In terms of translators…maybe those who may not be of the same culture or the way that they’re explaining the medical terminology may be intimidating” [53, p.6]. Medical mistrust was identified as negatively impacting vaccination intention [53,67], with trust in personal HCPs as sources of vaccine information positively associated with Covid-19 vaccination intention among Asian people [46]. Covid-19 vaccine-specific concerns arose around safety, adverse effects, and efficacy [53,64,67], including the need for population-specific evidence: “There has been a lot of concerns in my family on how the vaccine works for people with heart disease, which really affects…a lot of the Filipino community, and also those with respiratory diseases” [53, p.6].

#### American Indians

No studies among those reviewed focused on Covid-19 vaccination among American Indians or Alaskan Natives; one source provided data on American Indians [53]. Structural factors arose regarding distrust of government, desire for autonomy (“to be respected)”, challenges in access to digital technology and the internet, lack of quality translations and tailored information, lack of employment benefits, insurance, and transportation to get vaccinated [53]. In addition to distrust ascribed to the politicization of vaccination, a participant decried structural-level challenges for elders: “Some elders…they’re houseless…there’s no cell phone, they ride public transportation. How is it going to get distributed [to them]…the most vulnerable… it feels like our culture always gets the shitty end of the stick” [53, p.6]. Covid-19 vaccine-specific concerns arose around side effects, efficacy, and multiple doses—associated with challenges for Native families due to the need for several trips to vaccination sites.

#### LGBTQ+ people

Two sources focused on Covid-19 vaccination among LGBTQ+ people [83,86]. At the structural level, medical mistrust based on historical discrimination against sexual and gender minorities was negatively associated with Covid-19 vaccination intentions [86]. Loss of employment and housing insecurity were described as factors that underscored the threat of the pandemic among LGBTQ+ people, thus motivating vaccination. At the social and community level, altruistic motivations were positively associated, while anticipated stigma was negatively associated with vaccination intentions [83,86]. Individual-level factors impacting vaccination were described in perceived threat of Covid-19: “negative experiences, along with the reported high levels of perception of the seriousness of Covid-19 infection, are likely driving the high levels of willingness to be vaccinated” [83,p.9]. Covid-19 vaccine-specific concerns included adverse effects and safety [86].

#### People with disabilities

Nine articles focused on challenges in Covid-19 vaccination for people with disabilities (PWD) [12,57,58,64,67,72,76,88,91]. At the structural level, five studies addressed the impact of trust in government: three reported a positive association with Covid-19 vaccination [57,58,88], while two described governmental distrust contributing to VH [67,72]. Strict public health measures and widespread reliance on digital technology during the pandemic posed structural barriers in access to Covid-19 vaccination for PWD [12,72,76,88]. While PWD were described as more likely than people without a disability to endorse Covid-19 vaccination, they were more likely to report difficulties booking online vaccination appointments and locating and travelling to vaccination sites [12].

At the social and community level, having a trusted HCP or clinic exerted a positive influence on Covid-19 vaccination [12,57,76,88,91]. For example, among people living with multiple sclerosis (MS), MS-specialist HCPs and the National MS Society were deemed trusted sources: “well-positioned to provide accurate education about Covid-19 disease and vaccine considerations…specific to the MS population as well as their individual patients” [57,p.3].

At the individual level, perceived threat of Covid-19 was a positive correlate of Covid-19 vaccination among PWD, including individuals with chronic diseases and multiple comorbidities [12,57,58,64,72,76,91]. Covid-19 vaccine-specific concerns included adverse effects, speed of vaccine development, vaccine safety, and efficacy [58,64,67,72,76,88,91]. In addition to “safety and efficacy…the top Covid-19 vaccination concerns”, apprehension arose around “the effect of the vaccine on MS symptoms and implications for [immunosuppressive] DMT [disease-modifying therapy] use” [88, p.1078].

### Justice-involved people

Four articles focused on Covid-19 vaccination among justice-involved (JI) individuals (i.e., currently or formerly convicted or incarcerated) [54,63,66,84]. At the structural level, medical mistrust and distrust of government exerted a negative impact on Covid-19 vaccination among individuals in detention facilities [54,63,84] “based on their interactions with law enforcement or the justice system or their experiences with institutional racism” [84, p.476]. A study with incarcerated women described “a direct correlation between trust in the pharmaceutical industry and willingness to receive the vaccine”: “I’ve watched too many people die because pharmaceuticals… So I don’t know. Pharmaceutical companies just see a lot of big money in there” [63, p.895]. One study described mitigation of structural-level barriers to Covid-19 vaccination through tailored vaccine education and “early access to vaccine through a dedicated federal allocation” for incarcerated individuals and facility staff [66,p.5889].

At the social and community level, family and community influence was associated with Covid-19 VH [54,63,66] among JI individuals in the absence of tailored public health messaging: “The probable low health and informational literacy of these women leave them vulnerable to influence from family and friends, many sharing and promoting disinformation and conspiracy theories frequently shared by homegrown anti-vaccine organizations” [63, p.895]. Social proofing exerted a positive influence: “some people may have been encouraged to accept vaccination because they could see their peers being vaccinated” [54,66, p.5889].

At the individual level, all four studies reported low perceived risk of contracting Covid-19 as a correlate of VH among JI people [54,63,66,84]. A study in correctional and detention facilities in 4 states reported that of participants (n=1823/5110) who would refuse Covid-19 vaccination, 18.9% (n=344/1823) “did not perceive themselves to be at risk for Covid-19 or thought vaccination was unnecessary” [84, p.475]. Covid-19 vaccine-specific concerns arose around vaccine safety and efficacy [54,63,84], which was associated with “lack of access to reliable information” [54, p.375].

#### People living with HIV/AIDS

Two articles focused on factors impacting Covid-19 vaccination intentions among PLHA [70,80], with two additional sources providing PLHA-specific data [51,83]. At the structural level, medical mistrust and distrust of government among PLHA were associated with lower Covid-19 vaccination intentions [51,70,80]. A study with HIV-positive and predominantly sexual minority Black Americans (n=101) indicated that “The most prevalent general mistrust beliefs (endorsed by about half or more than half) concerned withholding information or a lack of honesty by the government,” which was correlated with VH [51, p.203]. A study with HIV-positive Black and Latinx Americans described unemployment being exacerbated by the pandemic, with negative impacts on access to healthcare and Covid-19 vaccination [70]. At the social and community level, the positive influence of personal HCPs as trusted sources of information on Covid-19 vaccination was reported [51,70,80]: “Lower hesitancy scores were present in participants who reported that they would get vaccinated if their provider recommended it” [80, p.98].

At the individual level, perceived threat was associated with Covid-19 vaccination intentions: “…HIV-positive men may have more concerns about the seriousness and perceived impact of Covid-19 infection on their health” [80, 83, p.9]. Covid-19 vaccine-specific concerns arose around vaccine safety, adverse effects, and efficacy [51,70,80], with some PLHA concerned that their health condition may make Covid-19 vaccination unsafe [80].

#### People who use drugs

Two studies focused on Covid-19 vaccination among people who use drugs (PWUD) [73,92], with one additional source providing disaggregated data on PWUD [64]. At the structural level, barriers to accessing healthcare and Covid-19 vaccines, as well as medical mistrust and distrust in government were associated with lower Covid-19 vaccination intentions [64,73]. One study noted vulnerability to misinformation among PWUD amid medical mistrust: “As a marginalized population, patients with illicit drug use are often detached from and mistrust the healthcare system—by extension, these patients may be more reliant on illegitimate information sources” [64, p.803]. At the individual level, mask wearing was associated with vaccine trust, both described as a function of accurate Covid-19 information [73]. Covid-19 vaccine-specific factors arose around safety and efficacy concerns that may negatively impact uptake [73].

#### Immigrants and refugees

One study focused on Covid-19 vaccination among immigrants and refugees [93], with one additional source providing disaggregated data [61]. At the structural level, barriers to healthcare and Covid-19 vaccine access were described as negatively impacting vaccination [61,93]. A study with Latinx immigrant families identified “fear of encountering law enforcement/immigration as barriers to access health care” [61, p.750]. Limited access to information in one’s preferred language, lack of reliable information from trusted sources, constraints in access to the internet and digital technology, and lack of transportation and employment benefits (i.e., paid sick days) were identified as barriers to Covid-19 vaccination [93]. At the social and community level, family influence was demonstrated in a national online survey of refugees (n=435): among those who reported intention to receive a Covid-19 vaccine, nearly two-thirds (65.0%; n=199/306) indicated that it was motivated in part to protect their family members [93]. Covid-19 vaccine-specific concerns about vaccine safety, adverse effects, and efficacy were associated with VH, with two-thirds (71.3%; n=92/129) of refugees who reported no intention or being unsure of accepting Covid-19 vaccination indicating concerns about adverse effects [93].

#### People experiencing homelessness

One article focused on challenges to Covid-19 vaccination for people experiencing homelessness [71]. At the structural level, Covid-19 VH was described as a function of “a more general distrust of systems (e.g., health and homeless services) that are not designated to meet their needs” [71, p.6]. Trusting the information received from friends, family, and social media rather than “official news sources” was associated with greater Covid-19 VH [71]. At the individual level, general vaccine aversion and low Covid-19 risk perception, and vaccine-specific concerns about safety, adverse effects, and efficacy were associated with VH among people experiencing homelessness [71].

## Discussion

This scoping review of Covid-19 vaccine hesitancy and broader underutilization of Covid-19 vaccination provides evidence of multilevel determinants of delays or refusal of Covid-19 vaccination across marginalized populations in the U.S. Overall, a preponderance of structural-level factors were identified that impact Covid-19 vaccination across marginalized populations. Logistical and economic constraints including lack of transportation to distant locales, lack of paid sick days, lack of availability of vaccination at familiar/local community venues, and barriers in access due to being unemployed, unstably housed, uninsured/underinsured, and the digital divide (i.e., access to digital technology and the internet, and technical and financial ability to utilize it [94]) were compounded by lack of culturally and linguistically appropriate information and access to trusted HCPs. Collectively, these findings suggest that approaches to undervaccination among marginalized populations that focus predominantly on individual-level, psychological factors may fail to capture the breadth and impact of determinants of Covid-19 vaccination.

Importantly, we identified evidence for differential impacts of medical mistrust, as a structural factor [29], and trust in a personal HCP or clinic, as a social/community factor. The influence of medical mistrust reported with respect to government public health, healthcare systems, and the pharmaceutical industry, emerged across multiple marginalized populations— including African American, Latinx, LGBTQ+ populations, and their demographic intersections—attributed in part to historical discrimination and past unethical medical research and interventions, as well as present-day discrimination in healthcare and government institutions. Nevertheless, having a trusted HCP, particularly with shared race or ethnicity [26,51,69,70,74,82,85], or a familiar source of primary healthcare, mitigated Covid-19 VH— despite broader mistrust of healthcare systems and government public health. JI populations, PEH, and PWUD expressed medical mistrust due to generally negative experiences with government and healthcare institutions. However, as in the case of JI populations, proactive interventions to promote tailored education along with policy decisions to prioritize Covid-19 vaccination in prisons served to mitigate VH and promote vaccination [66].

Personal HCPs and usual care venues were widely reported to assuage concerns and reduce VH, indicating a crucial role for primary healthcare [95]. In this light, the absence of information from trusted sources tailored to address healthcare needs experienced as most relevant to one’s own community—reported among Black and Pilipino populations, PLHA and PWD—and the lack of availability of Covid-19 and vaccine information in one’s primary language (reported among Latinx, Asian, and immigrant/refugee populations) emerged as substantial missed opportunities to promote Covid-19 vaccination. Access to Covid-19 vaccination from a known HCP or clinic may be particularly important for marginalized communities, who are more predisposed towards medical mistrust due to negative interactions with the healthcare system and other societal institutions, and more vulnerable to discrimination in healthcare [96,97].

Overall, the conflation of structural factors—documented in multifaceted barriers in access to Covid-19 vaccination and systemic mistrust of healthcare systems—with individual and social/community factors, such as perceived threat of Covid-19, perceived risk of transmission, general vaccine attitudes, endorsement from one’s personal HCP, and family influence, each of which may impact on “decisional conflict” more specifically aligned with VH [32], may fail to foster solutions that effectively address the identified problems [34,95,98]. Moreover, the attribution of structural-level barriers to individual volition and deficits may exacerbate alienation and mistrust among populations already marginalized due to systemic discrimination, stigma, and adverse SDOH [96].

Interestingly, some of the same ostensible factors showed negative or positive associations with VH depending on the population and context. For example, family influence exerted a positive impact on Covid-19 vaccination among Latinx communities and immigrant/refugee populations through familial sharing of health information and desire to protect one’s family. However, among other populations, the context of medical mistrust and the virtual vacuum created by the absence of access to culturally tailored public health information rendered family influence a liability, resulting in promulgation of conspiracy theories and mistrust of Covid-19 vaccines among some African Americans [81], PEH [71], and JI persons [63]. These examples underscore the importance of approaching VH as a population-specific and context-dependent phenomenon [6,7]—in addition to disentangling the assessment of systemic factors from individual-level and social/community concerns [99]. Consequently, the same intervention applied to what might be understood as the same factor may exert opposite effects on Covid-19 VH across different populations.

Finally, facilitators of Covid-19 vaccination emerged despite constraints. For example, findings in common across PLHA and PWD—among whom one might reasonably expect heightened concerns about vaccination due to underlying comorbidities and potential interactions with routine medications—included the benefits of regular contact with a personal HCP or clinic; this promoted trust in the vaccine information received and confidence in the HCP’s ability to address population-specific concerns. Furthermore, intersectional strengths reported regarding Latinx immigrant and refugee populations included motivations to protect one’s community and cultural norms around familial sharing of health information, which served to mitigate VH. Beyond a deficit model, community assets and sociocultural facilitators may be leveraged [100] to support Covid-19 vaccination.

Findings from this scoping review should be understood in the context of limitations. First, as a scoping review, our objective was to map and synthesize evidence on Covid-19 VH and undervaccination among marginalized populations; we cannot draw causal conclusions about the associations identified, nor was our aim to quantify Covid-19 vaccination intentions or uptake, more apropos of meta-analysis. However, our scoping review methods reveal nuanced information through synthesis of quantitative and qualitative findings on population-specific barriers to Covid-19 vaccination and suggest important directions for future research. Second, additional populations to those included might be characterized by marginalization, although we based our focal populations on previous evidence and a priori criteria [42,43]. Third, while our initial search comprised studies from the U.S. and Canada, no studies were identified from Canada that met inclusion criteria during the specified time frame, possibly due to the later availability of Covid-19 vaccination to the general adult population [41]. Studies published after the time frame of our review might include additional populations, data, and study locales, although we purposely focused on the initial introduction of Covid-19 vaccines before subsequent booster doses and bivalent vaccines. Lastly, factors associated with VH and underutilization of monovalent and bivalent boosters may differ from those revealed for initial Covid-19 vaccination, although vaccine inequities remain [23].

In conclusion, we identified multilevel factors that influence Covid-19 VH and broader underutilization of Covid-19 vaccination, several of which may apply to other vaccinations. This scoping review suggests that distinguishing among individual and social/community factors that may fuel decisional ambivalence characteristic of Covid-19 VH—”a state of indecision and uncertainly about vaccination before a decision is made to act (or not act)” [35, p.58]—and factors attributable to systemic and structural barriers in access to vaccination and historically-justified mistrust of healthcare institutions, government public health, and the pharmaceutical industry, may support evidence informed strategies to promote Covid-19 vaccination among marginalized communities. Nevertheless, distilling and applying lessons from Covid-19 vaccination to effectively and equitably address VH and broader challenges in vaccine access may be contingent on resourcing and scaling up community partnerships and local health infrastructures [23,25,96,100], national population-specific public health initiatives to promote vaccination [101,102], and structural reforms beyond those traditionally associated with public health.

## Data Availability

All relevant data are within the manuscript and its Supporting Information files.

## Acknowledgements

We would like to thank Nalini Singh, University of Toronto Research Librarian, for assistance in designing the literature search strategy and Dares Wotansirigul for uploading sources into Covidence software.

## Funding

This work was supported by the Canadian Institutes of Health Research [GA3-177731].

## Declaration of competing interest

The authors declare the following financial interests/personal relationships which may be considered as potential competing interests: Peter A. Newman is a member of the Editorial Board of PLOS ONE.

## Supporting information

**S1 Table. Sample search string**

## Notes

### Clinical Protocols

https://journals.plos.org/plosone/article?id=10.1371/journal.pone.0266120

### Funding Statement

This work was supported by the Canadian Institutes of Health Research [GA3-177731, recipient: PAN].

### Author Declarations

This review did not include any human subjects or data collection and thus did not require ethical approval.

